# Evaluation of determinants of the serological response to the split-inactivated influenza vaccine

**DOI:** 10.1101/2021.10.07.21264416

**Authors:** Shaohuan Wu, Ted M. Ross, Michael A. Carlock, Elodie Ghedin, Hyungwon Choi, Christine Vogel

## Abstract

The seasonal influenza vaccine is only effective in half of the vaccinated population. To identify determinants of vaccine efficacy, we used data from >1,300 vaccination events to predict the response to vaccination measured as seroconversion as well as hemagglutination inhibition (HAI) levels one year after. We evaluated the predictive capabilities of age, body mass index (BMI), sex, race, comorbidities, prevaccination history, and baseline HAI titers, as well as vaccination month and vaccine dose in multiple linear regression models. The models predicted the categorical response for >75% of the cases in all subsets with one exception. Prior vaccination, baseline titer level, and age were the strongest determinants on seroconversion, all of which had negative effects. Further, we identified a gender effect in older participants, and an effect of vaccination month. BMI played a surprisingly small role, likely due to its correlation with age. Comorbidities, vaccine dose, and race had negligible effects. Our models can generate a new seroconversion score that is corrected for the impact of these factors which can facilitate future biomarker identification.

## Introduction

Influenza virus infections represent a continuous threat to public health, as vaccine effectiveness is typically low, ranging from 19% to 60% during the 2009 to 2018 seasons in the United States, according to the Center for Disease Control (https://www.cdc.gov/flu). The widely used split-inactivated influenza vaccine is typically quadrivalent with an antigen for all four virus subtypes, H1N1 and H3N2 (Influenza A) subtypes, and Yamagata and Victoria lineages (Influenza B). Understanding predictors of vaccine efficacy is an ongoing public health challenge.

While antigenic drift and shift caused by frequent mutations in circulating viral strains are long-known influencers of vaccine efficacy, there is increasing recognition of other factors intrinsic to the human host which impact vaccine efficacy and/or severity of an influenza infection. These factors can be genetic (Orrù *et al*, 2013; Brodin *et al*, 2015; Franco *et al*, 2013), epigenetic (Zimmermann *et al*, 2016), or represent pre-existing immunity (Henn *et al*, 2013; HIPC-CHI Signatures Project Team & HIPC-I Consortium, 2017; Voth *et al*, 1966; Beyer *et al*, 1996) as caused by prior infection or vaccination (Sung *et al*, 2021; Gouma *et al*, 2020; Zost *et al*, 2017). Further, demographic factors such as age (Henry *et al*, 2019; Henry *et al*, 2019; Goodwin *et al*, 2006), obesity (Honce & Schultz-Cherry, 2019; Honce *et al*, 2020), and sex (Klein & Flanagan, 2016; Fink *et al*, 2018; Voigt *et al*, 2019) are thought to play a role. In addition, recent studies have shown that vaccination time during a flu season can also affect the response to the vaccine (Penkert *et al*, 2021). However, many of these factors are intercorrelated, e.g. prior vaccination and baseline antibody titer level as well as age, obesity, and other comorbidities. In addition, most existing studies examined the effect of one or a few of these factors (Penkert *et al*, 2021; Sung *et al*, 2021; Gouma *et al*, 2020; Goodwin *et al*, 2006) without comprehensive integration.

Here, we addressed this gap in knowledge by using a large cohort dataset to construct multiple linear regression models that predicted the response to the split-inactivated influenza vaccine based on nine variables known for participants. We performed the prediction separately for three different age groups, as well as for seroconversion after 3-4 weeks and antibody titer levels in the subsequent year. We evaluated the impact of each individual factor while controlling for the effects of the remaining factors.

## Results

### Mining a large cohort study with >1,300 vaccination events

To construct the models, we used one of the largest cohort studies available, involving ∼700 participants monitored over five flu seasons (cohorts), producing 1,368 vaccination events (**Figure 1**). We predicted both seroconversion (*Seroconversion*), measured as the log_2_ ratio between HAI (hemagglutination inhibition) titer levels against the four vaccine strains 3 or 4 weeks post vaccination (D21 or D28, respectively) and HAI titer at D0, and the baseline (BL, D0) HAI titer levels in the subsequent year (*BaselineSY*). In addition to demographic factors, such as age, body mass index (BMI), sex, race, comorbidities, and prevaccination history of participants, we included month of vaccination (September to February), and vaccine dose (for participants >=65 years old) to predict *Seroconversion* and *BaselineSY*. Based on the longevity of the vaccine’s effect (**Supplementary Figure S1**), we defined participants as prevaccinated if they had received the vaccine in the previous year, and as naive if they had had no vaccine within the last three years, removing cases with a mixed status. We also used HAI levels at D0 or D28 for prediction of *Seroconversion* and *BaselineSY*, respectively.

**Figure 1.**
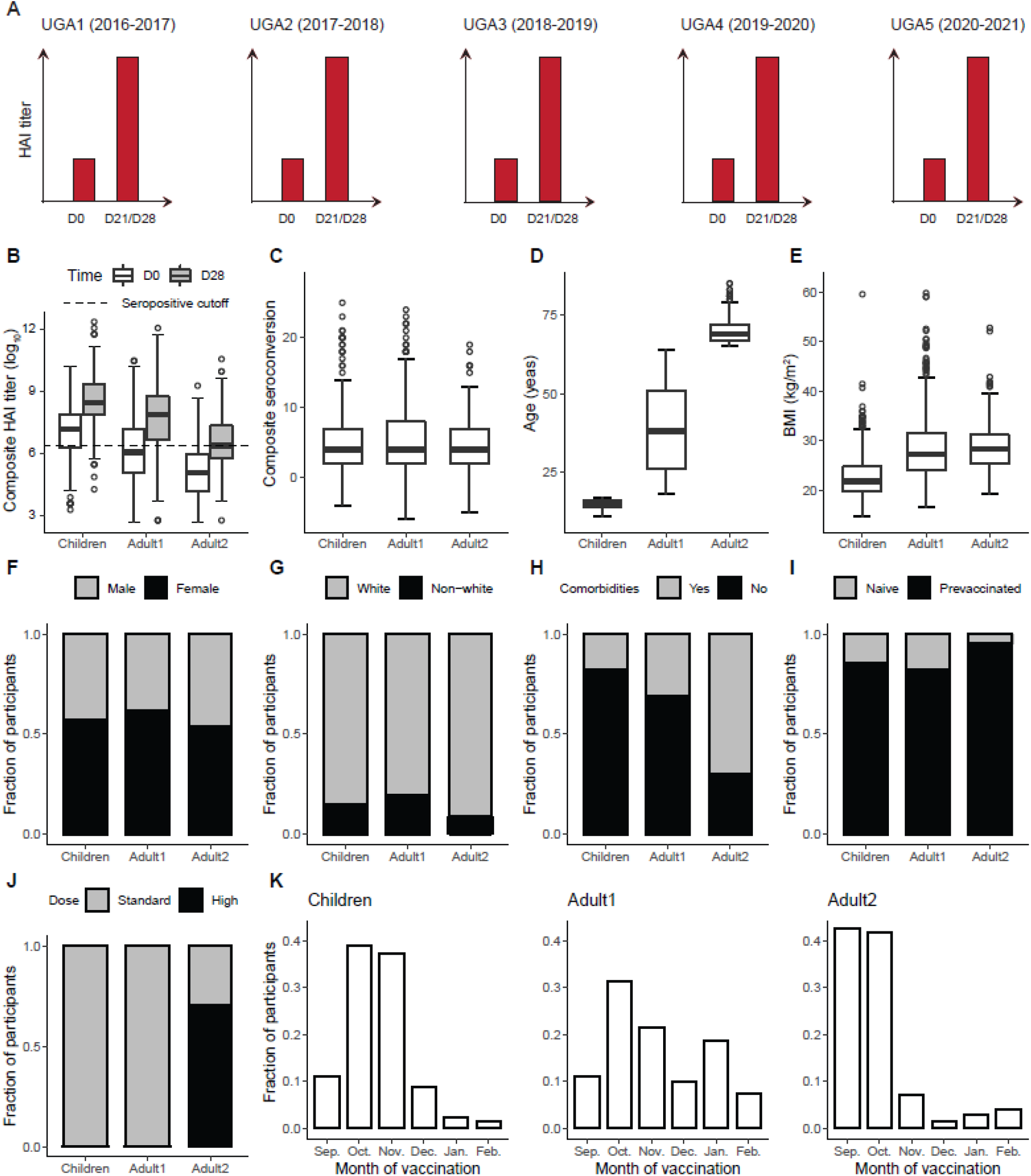
The UGA flu vaccination cohort includes diverse participants with multi-dimensional prior information. **A**. Data for flu vaccine cohort by University of Georgia at Athens (UGA) spanning 5 flu seasons. For each of the human participants, day 0 (D0) and day 21/28 (D21/D28) antibody titer levels against each of four vaccine strains were measured using the hemagglutination inhibition assay (HAI). **B**. Distribution of composite D0 and D28 HAI titer levels in the Children (<18 years old), Adult1 (18-64 years old), and Adult2 (>=65 years old) subpopulations. Similar to the definition of composite seroconversion, composite D0/D28 titer is defined as the sum of log_2_ (D0 titer level) or (D28 titer level) across 4 vaccine strains. Seropositivity cutoff is the composite titer level when titer is 40 in all 4 strains (4*log_2_ (40)). HAI, hemagglutination inhibition assay. **C-E**. Distribution of composite seroconversion, age, and BMI in 3 subpopulations. **F-J**. Distribution of categorical variables in 3 subpopulations. For comorbidities variable, *yes* indicates having >=1 of the comorbidities that are surveyed and *no* indicates having none (**H**). For vaccine dose, high dose is offered as an option only to Adult2 subpopulation (**J**). **K**. Fraction of participants that are vaccinated in each month in a flu season. Sep., September; Oct., October; Nov., November; Dec., December; Jan., January; Feb., February.

We separated participants into three subpopulations: *Children* (12-17 years), *Adult1* (18-64 years), and *Adult2* (>=65 years). **Figure 1** shows the distributions of variables across these three groups. Most participants across all groups were prevaccinated which is a substantial determinant of the baseline HAI levels (**Figure 2**). However, despite prior vaccination, the average baseline HAI level was much lower in the *Adult2* group compared to the other two groups: <25% of *Adult2* participants were seropositive at D0, the beginning of the respective study (**Figure 1**). Notably, the three subpopulations exhibited a similar seroconversion after the vaccination, indicating that other factors were in play besides baseline level (**Figure 1**).

**Figure 2.**
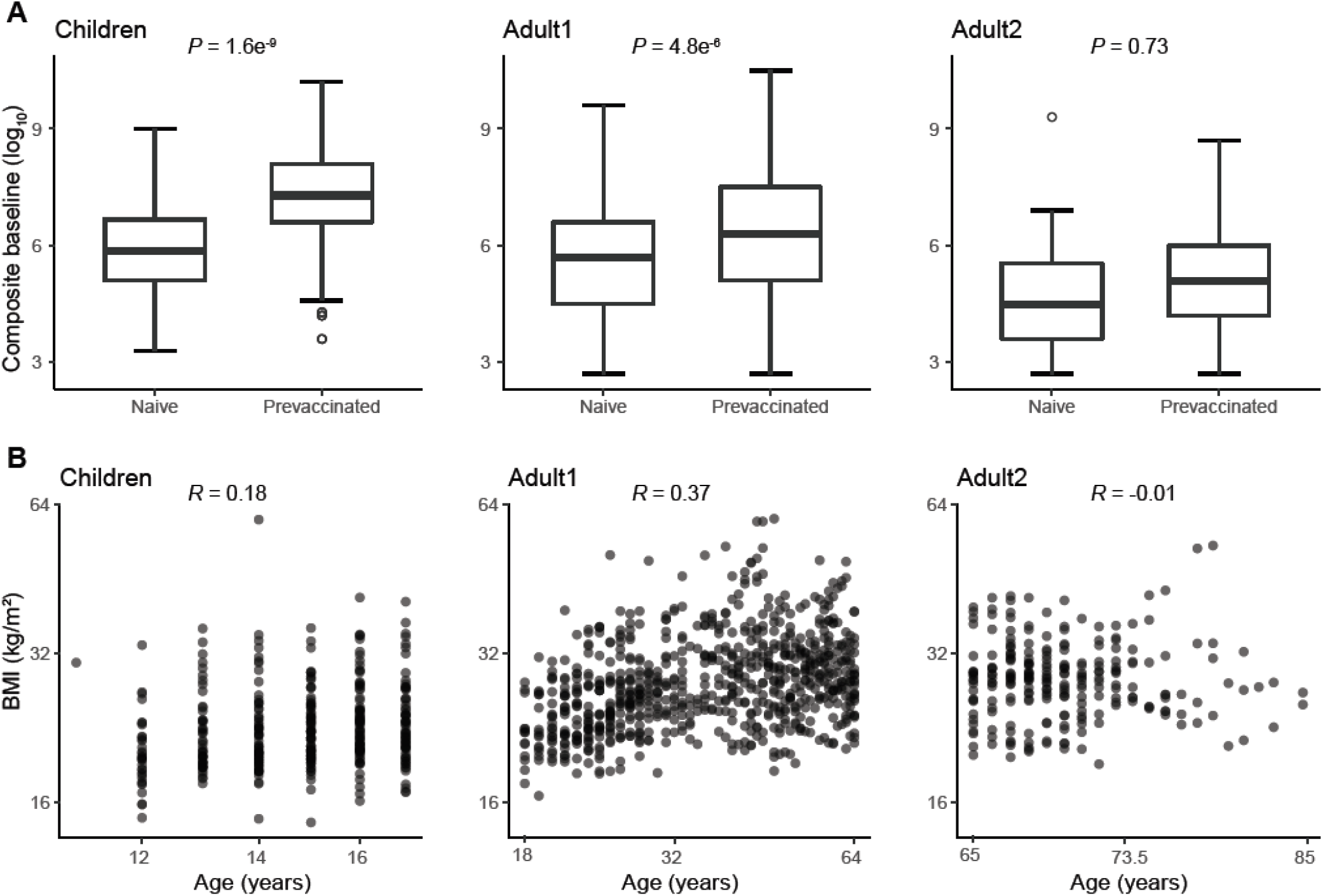
Some priors are intercorrelated. **A**. Correlation between prevaccination history and baseline across three subpopulations. *P* value is calculated from the *T* test. **B**. Correlation between age and BMI across three subpopulations. *R* is calculated from Pearson’s correlation.

Other variables showed different distributions across three subpopulations, which, in part, impacted their ability to predict the outcome of the vaccination. As expected, *Adult1* and *Adult2* had on average higher BMIs than *Children*, and most *Adult2* participants had one or more comorbidities (**Figure 1**). The three subpopulations were similar with respect to distributions of the remaining demographic factors, i.e. gender and race (**Figure 1**). Only *Adult2* participants had been offered the high dose of the vaccine (**Figure 1**). Finally, *Children* and *Adult2* participants had primarily been vaccinated in the first three months of a flu season while *Adult1* participants were vaccinated evenly throughout the season (**Figure 1**). For that reason, dose and vaccination month were only useful predictors in the *Adult2* and *Adult1* subpopulations, respectively (see below).

### Predicting over three quarters of the vaccine response

We predicted *Seroconversion* and *BaselineSY* for both the four vaccine strains individually and the composite value calculated as the sum of the values across four strains (**Figure 3**). For the prediction of *Seroconversion*, we trained the models on four UGA cohorts and evaluated the prediction on the fifth, independent cohort. For the prediction of *BaselineSY*, we trained the models on three of the four newly assembled ‘BL cohorts’ (**Methods**) and evaluated the prediction on a fourth, independent BL cohort (**Figure 3**).

**Figure 3.**
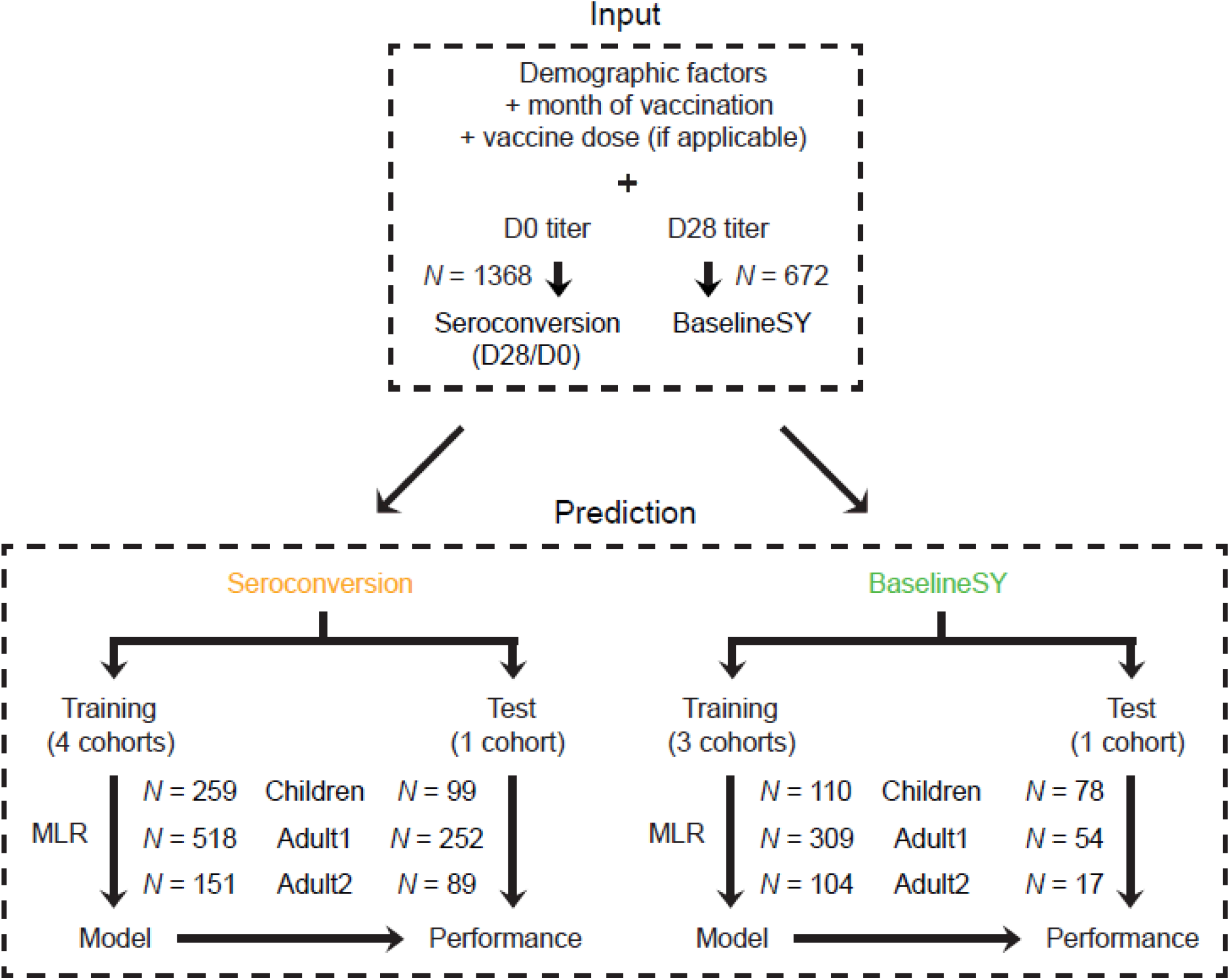
A multiple linear regression model aims at removing confounding factors when predicting the response to vaccination. Variables for *Seroconversion* prediction and *BaselineSY* prediction, respectively, in each of the 3 subpopulations, and machine learning strategy used to predict *Seroconversion* and *BaselineSY*. MLR, multiple linear regression.

First, we evaluated the models with respect to prediction of categories (high vs. low/no *Seroconversion*; seropositive vs. negative *BaselineSY*) (**Figure 4**). All three subpopulations predicted *Seroconversion* and *BaselineSY* accurately at 74% or more in independent test sets. The models performed best for the subsequent year’s baseline HAI levels for *Adult2* subpopulation (>=65 years old) (94% correct predictions). The models performed worst for prediction of *Seroconversion* and *BaselineSY* for the *Adult1* subpopulation (18-64 years old), indicating that for these participants, additional factors, e.g., specific comorbidities, impacted the response to vaccination. **Supplementary Figure S3** shows the results when evaluating the models across the entire dataset.

**Figure 4.**
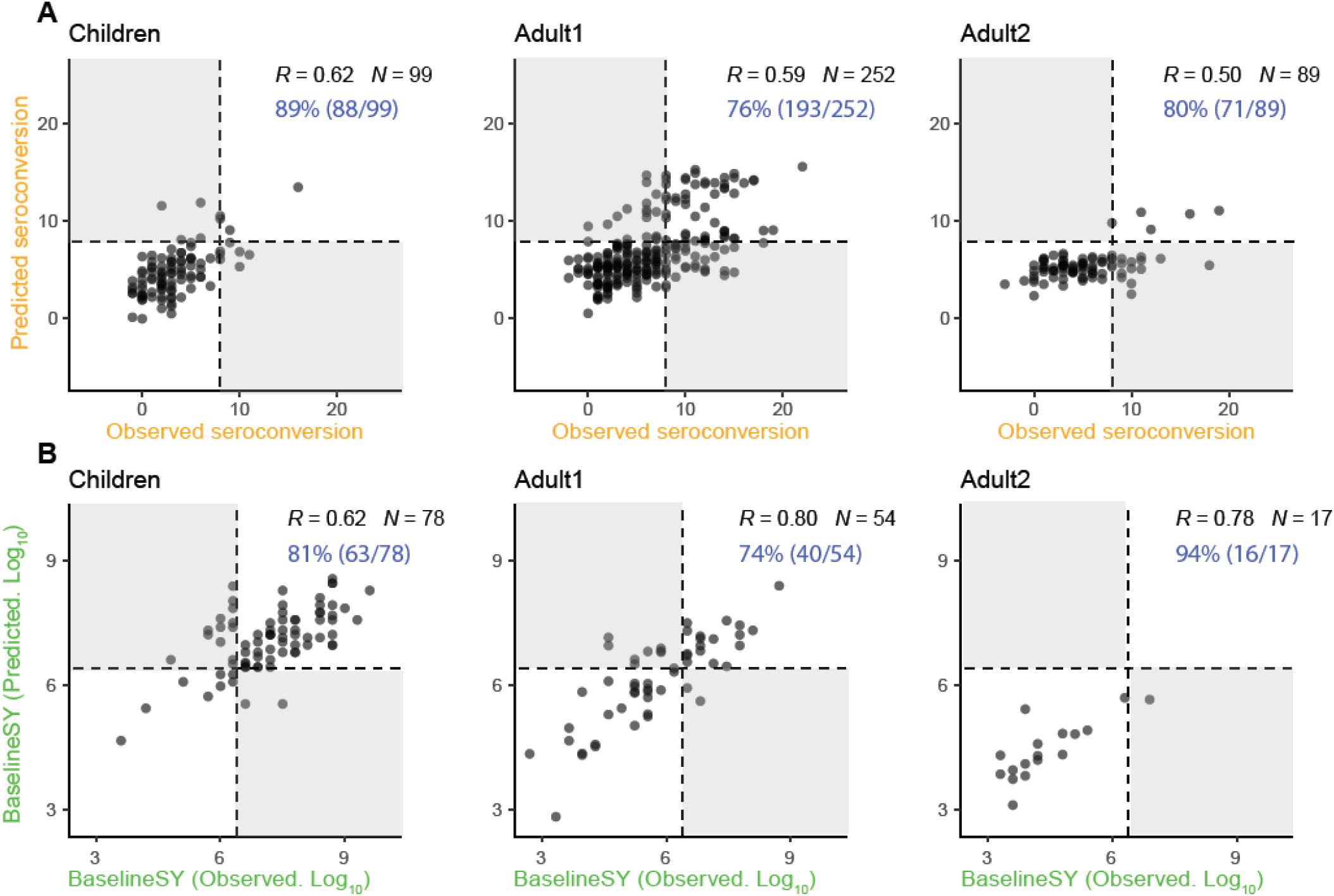
The model predicts seroconversion and long-term titer levels with high accuracy. **A**. Observed vs. predicted values for *Seroconversion*. Predicted values are obtained for the test set only (UGA4, see **Figure 1**). Dashed lines mark the cutoff for high seroconversion (>=8). White and grey quadrants show areas of correct and incorrect prediction of high and low/no seroconversion, respectively. Percentages listed in blue denote correct predictions. **B**. Observed vs. predicted values for *BaselineSY*. Predicted values are obtained for the test set only (repeated participants in cohorts 3-4). Dashed lines mark the cutoff for seropositivity (>=4*log_2_ (40)). White and grey quadrants show areas of correct and incorrect prediction of seropositive and seronegative baseline, respectively. Percentages listed in blue denote correct predictions.

### Identifying major predictors of *Seroconversion* and *BaselineSY*

Next, we estimated the relative importance of each variable in predicting *Seroconversion* and *BaselineSY* for both composite scores (**Figure 5**) and individual strains (**Figure 6**). Importantly, our results show the *independent* effect of each variable, even though several variables are intercorrelated (**Supplementary Figure S2**). **Supplementary Tables S1** and **S2** contain the original data for plotting **Figures 5** and **6**, respectively.

**Figure 5.**
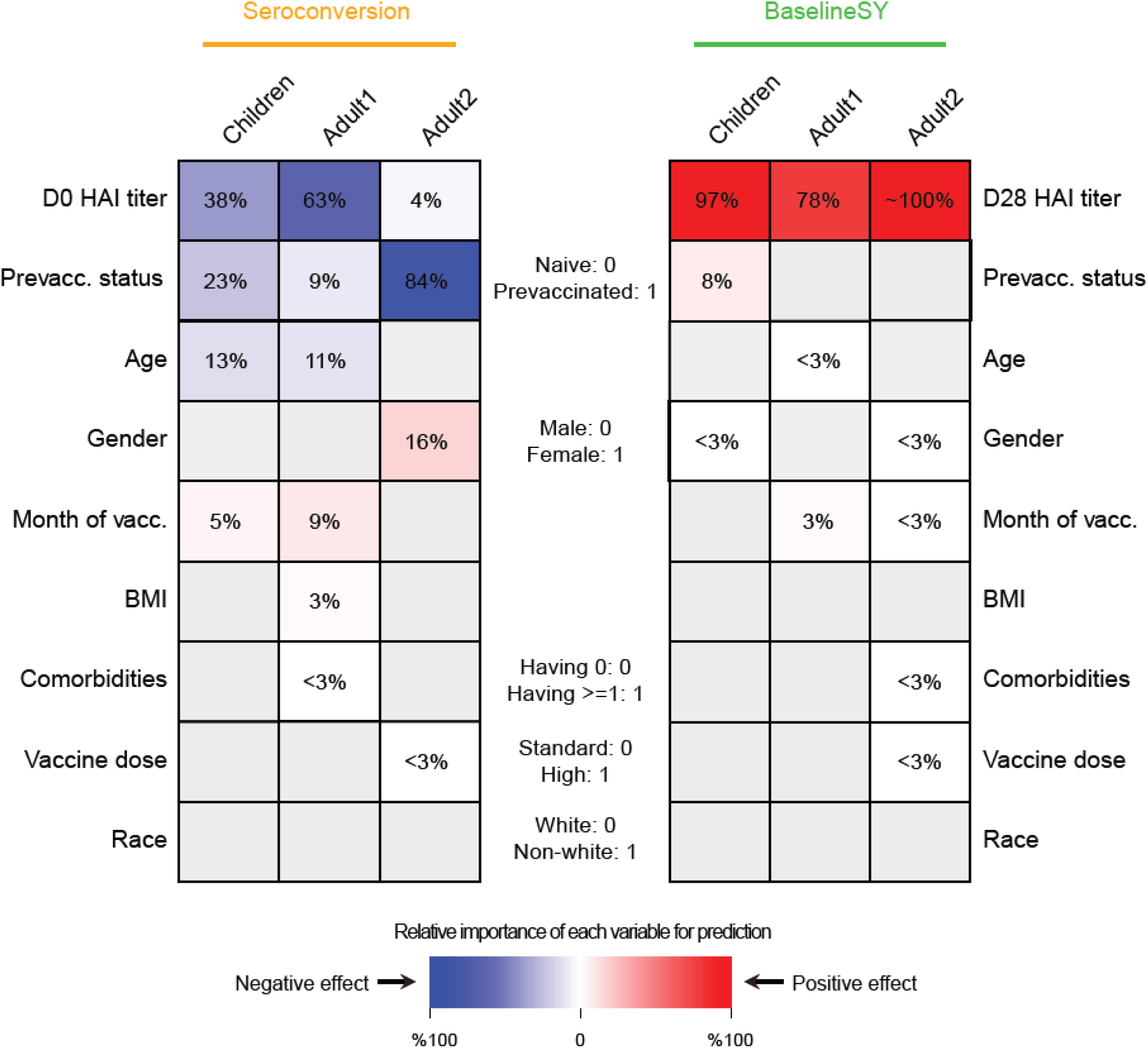
Individual priors have different contributions to response prediction. Relative importance of each variable for prediction. The intensity of the color in each cell denotes the importance, measured as percentage of drop in prediction accuracy measured as squared correlation (R^2^) between observed and predicted values (see **Methods**). Grey denotes non-selected variables. The color denotes the direction of the correlation between the variable and the predicted value, i.e. red and blue values denote positive and negative correlation, respectively, between the variable and *Seroconversion* or *BaselineSY*. For example, prevaccination status predicts both *Seroconversion* and *BaselineSY*; prevaccinated participants show lower (blue) *Seroconversion* (all subpopulations) and higher (red) *BaselineSY* (Children only). Variables with importance =< 3% or with a coefficient =< 0.1 in absolute value are labeled in white. HAI, hemagglutination inhibition assay; vacc., vaccination.

**Figure 6.**
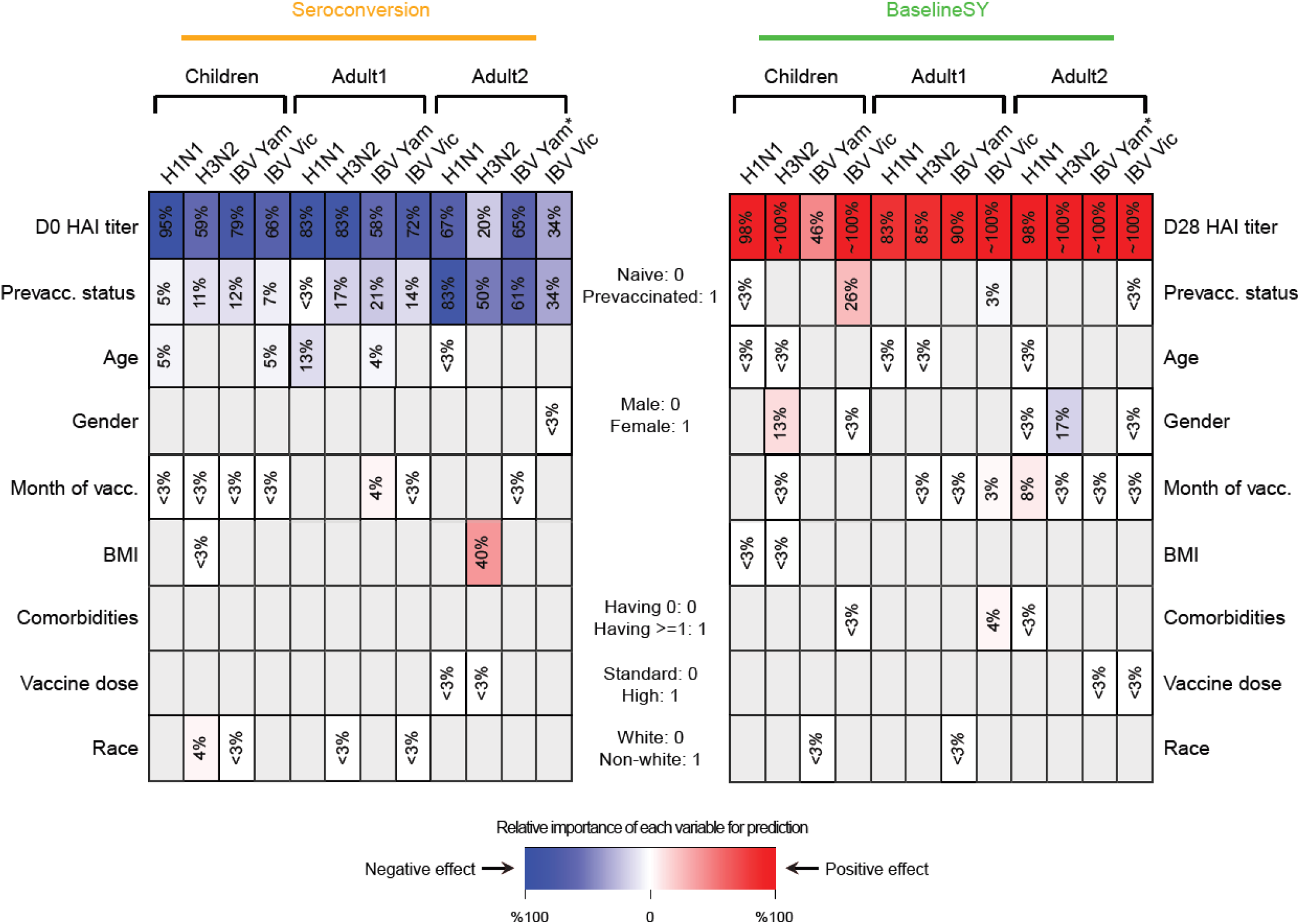
Strain-specific modelling reveals some detailed effects. Relative importance of each variable for prediction. The intensity of the color in each cell denotes the importance, measured as percentage of drop in prediction accuracy measured as squared correlation (R^2^) between observed and predicted values (see **Methods**). Grey denotes non-selected significant variables. The color denotes the direction of the correlation between the variable and the predicted value, i.e. red and blue values denote positive and negative correlation, respectively, between the variable and Seroconversion or BaselineSY. Variables with importance =< 3% or with a coefficient =< 0.1 in absolute value are labeled in white. IBV, Influenza B Virus; HAI, influenza hemagglutination inhibition assay; vacc., vaccination; Vic, Victoria; Yam, Yamagata. * IBV Yamagata strain is absent in the vaccine in UGA cohorts 1-4, but present in cohort 5.

As expected, prior vaccination and high HAI baseline levels predicted low *Seroconversion* and they explain the large majority of variation in each subpopulation. One exception was baseline HAI levels against the H3N2 strain in the *Adult2* subpopulation, in which the impact was negligible. Conversely, HAI titer levels at Day 21 or 28, but not prevaccination history, predicted most of the baseline titer level in the subsequent year (BaselineSY). The exception was *Children* in which prior vaccination had a small positive impact on the vaccine longevity.

Unexpectedly, older participants amongst the *Children* subpopulation also had lower *Seroconversion* than younger participants, similar to that among the *Adult1* subpopulation, which was largely driven by the effect of the H1N1 strain. This negative relationship was weak, but significant, as illustrated in **Figure 7** which depicts the relationship between the variable and the predicted value removing the effects of all other variables. This relationship suggests that immunosenescence contributable to natural influenza infections starts before adulthood. In comparison, participant age did not predict longevity of the antibodies which (as mentioned above) was almost exclusively determined by HAI titer levels.

**Figure 7.**
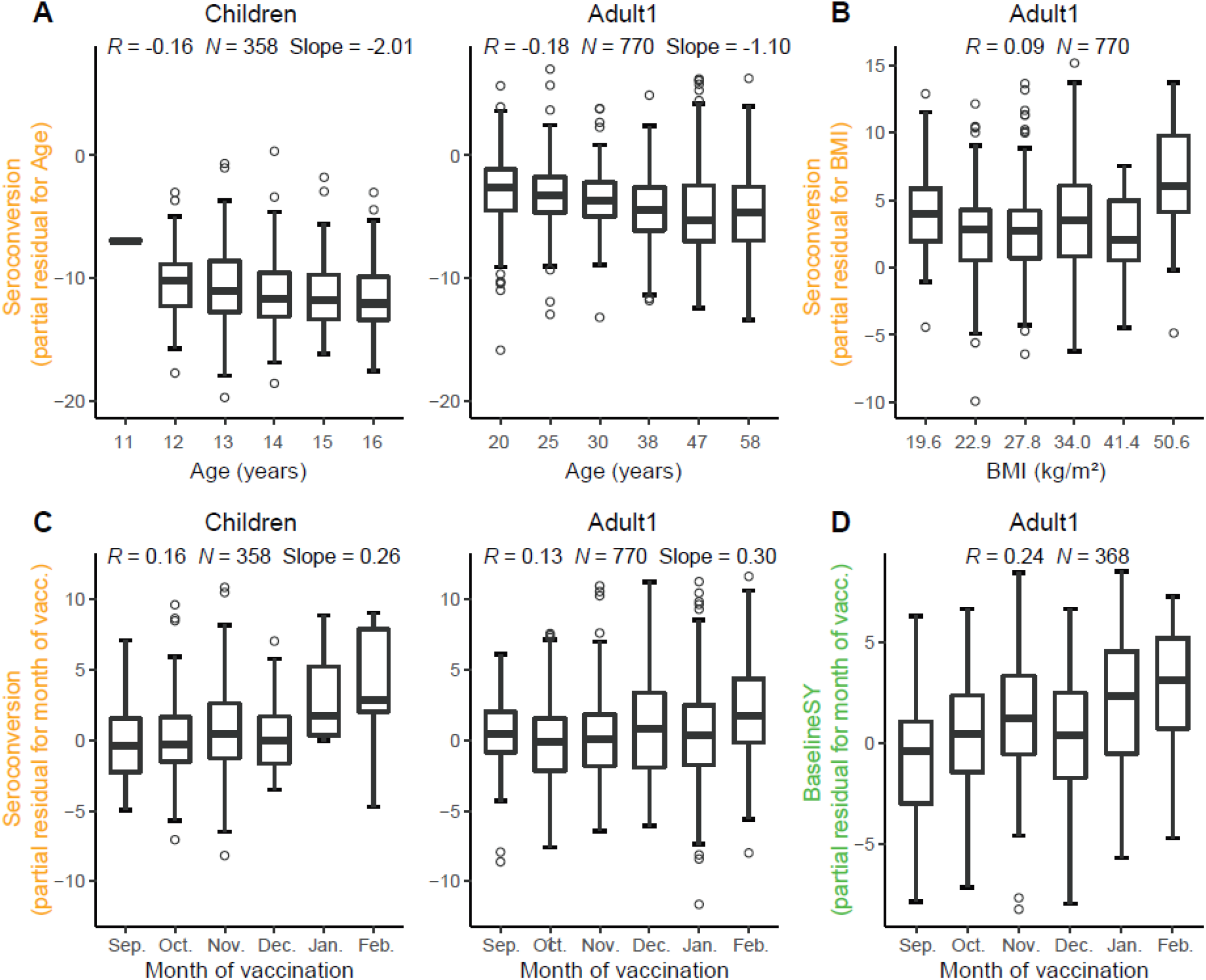
Relationships between age, BMI, and vaccination month, and the response to vaccination. Partial residual plots illustrating the relationship between a variable (x-axis) and the target value after removing the impact of all other variables (partial residual, y-axis). Target values are either *Seroconversion* (**A-C**) or *BaselineSY* (**D**). Vacc., vaccination. We grouped X-axes variables in **A-B** into 6 equally spaced bins, and showed the median for bin for better illustration, but we calculated *R* in all panels using the raw data.

Another major predictor was the month of vaccination: the later children and adults <65 years received the vaccine in a flu season, the higher *Seroconversion* was (**Figures 5** and **7**). For the *Adult1* subpopulation, this positive effect might have been mostly driven by the effect of the IBV Yamagata strain (**Figure 6**). The model could not evaluate this variable for the *Adult2* subpopulation due to the above-mentioned bias (**Figure 1**). The result is consistent with a recent finding (Penkert et al., 2021) and thought to link to natural influenza infections that occur more frequently later in the season, which could cause false positives if they occur during or shortly after the vaccination.

Surprisingly, body mass index had only a minor influence on composite vaccine efficacy: only in the *Adult1* subpopulation, BMI had a slightly positive effect on vaccine efficacy (**Figure 5** and **7**). When examining the influence of BMI on individual strains, BMI had a positive impact on the response to the H3N2 component in the *Adult2* subpopulation. BMI had no impact on vaccine longevity. These results appear to be in contrast to the known negative impact of obesity on influenza severity (Honce & Schultz-Cherry, 2019; Honce *et al*, 2020). We speculate that, since BMI and age are positively correlated (**Figure 2**), the strong age effect masks the effect of BMI on *Seroconversion*.

Further, we found that women had a higher response than men in the *Adult2* subpopulation, largely driven by the IBV Victoria strain (**Figures 5** and **6**). Race and comorbidities had no or only marginal effects on both *Seroconversion* and *BaselineSY*. The surprising lack of impact of comorbidities might be due to its intercorrelation with age (**Figure S2**), and the fact that comorbidities had only been noted as existing or not (i.e. present/absent) which provided little resolution of specific illnesses. Similarly, use of the high dose vaccine over the standard dose in the *Adult2* subpopulation had very little effect on vaccine efficacy.

## Discussion

We systematically examined the effects of nine factors on *Seroconversion*, i.e. the response to the vaccine 3-4 weeks after vaccination, and *BaselineSY*, i.e. the HAI titer levels in the subsequent year. To do so, we used a large cohort study with >1,300 vaccination events. We predicted *Seroconversion* and *BaselineSY* across three subpopulations based on participant age with an overall high accuracy: we predicted categorical classifications correctly in >3/4 of the cases and numerical values for >1/3 of the cases across most subpopulations (**Figure 4**).

Further, the models evaluated the contributions of the variables to the prediction of *Seroconversion and BaselineSY* (**Figures 5** and **6**). Across all age groups, the primary factors were vaccination status and baseline titer levels, followed by age and month of vaccination. These findings underscore the importance of pre-existing immunity, i.e. high baseline HAI titer levels against respective strains and longevity of the antibodies formed during prior infections or vaccinations. The impact of pre-existing immunity is an emerging concept (Boyd & Jackson, 2015; Boyd & Jackson, 2015; HIPC-CHI Signatures Project Team & HIPC-I Consortium, 2017; Gonzalez-Dias *et al*, 2020; Kotliarov *et al*, 2020; Tsang *et al*, 2020).

Importantly, we found that in all three subpopulations, prior vaccination and baseline titer levels had both shared and variable-specific contributions to *Seroconversion* (**Figures 2** and **5**). In adults, prior vaccination and/or baseline titer levels explained the large majority (if not all) of *Seroconversion*; the effect was smaller in *Children* perhaps due to a still maturing immune system. Further, the effect of prior vaccinations was visible for at least three years post vaccination (**Supplementary Figure S1**), indicating that antibody longevity is essential to consider when evaluating the vaccine response.

Similar approaches that corrected for baseline titer levels, such as producing a new score called adjMFC, have been very successful in identifying novel transcript markers of the vaccine response (Tsang *et al*, 2020; HIPC-CHI Signatures Project Team & HIPC-I Consortium, 2017). However, in contrast to adjMFC, our models included not only baseline titer levels, but a total of nine confounding variables. To our knowledge, our analysis is therefore the most comprehensive of its kind with respect to cohort size and number of variables modeled simultaneously, resulting in robust control of confounding effects and estimation of the effect size.

Our results are validated by previous findings. For example, two studies on small cohorts confirm the negative effect of prior vaccination on *Seroconversion* (Sung *et al*, 2021; Gouma *et al*, 2020). Further, a cross-cohort analysis confirms the negative effect of age on *Seroconversion* and seroprotection, i.e. the actual titer levels (Goodwin *et al*, 2006). While this analysis included data from 31 different cohort studies, it suffered from a less accurate estimation of the effect size of age, as several continuous variables had been treated as categorical. Further, our models show that participant sex affects *Seroconversion* in the *Adult 2* subpopulation, consistent with previous findings (Klein & Flanagan, 2016; Fink *et al*, 2018; Voigt *et al*, 2019). However, all of these prior studies examined only a single factor in its effect, therefore ignoring the contribution of other factors.

In comparison to the strong negative effect of age on *Seroconversion* in the *Children* and *Adult1* subpopulations, (**Figure 5** and **7**), age was not a predictor of the vaccine response amongst adults older than 65 years (*Adult2*), contradicting findings on increased flu severity amongst the eldery due to immunosenescence (Huang *et al*, 2019; Gounder & Boon, 2019). However, this result is not a contradiction but an extension of these previous findings: we showed that, indeed, participants older than 65 years have lower *Seroconversion* (**Figure 1**), but *within* this age group, age does not have any additional effect (**Figure 7**). The result indicates that the effect of immunosenescence perhaps reaches saturation beyond a specific age. Further, participants older than 65 years had the choice to receive the high-dose vaccine. However, in contrast to a previous study (DiazGranados *et al*, 2013), we did not find a positive effect of the high-dose vaccine (**Figure 5**).

Consistent with a recent finding examining the effect of time of vaccination on immune response (Penkert *et al*, 2021), we found that *Seroconversion* was higher when the vaccine was administered later in the season for *Children* and *Adult1* subpopulations. As most participants in the *Adult2* subpopulation received the vaccine in September or October of the season, we could not evaluate the effect of vaccination time in this group.

Our results also contribute to the discussion on the role of obesity in infection and vaccination. While obesity is a strong predictor of flu severity (Honce & Schultz-Cherry, 2019; Honce *et al*, 2020), conflicting findings exist on its effect on the response to vaccination. In response to an inactivated trivalent influenza vaccine, obese individuals have higher seroconversion than normal weight individuals, but their antibodies are shorter lived (Sheridan *et al*, 2012). In contrast, other studies found obesity to impact seroconversion negatively in both young and old individuals (Frasca *et al, 2016)*, or to have no influence on the response in the elderly or in children (Talbot *et al*, 2012; Callahan *et al*, 2014).

We hypothesize that these inconsistencies may arise from possible shortcomings in using BMI as a measure of metabolic health (Nuttall, 2015), as well as the mixed effect of confounding factors, primarily age. In light of these complex relationships, we interpret our results as follows: In *Children*, BMI varies relatively little (**Figure 1**) and is slightly positively correlated with age (**Figure 2**). The absence of a BMI effect might therefore be due to the predominant effects of prior vaccination, baseline titers and age (**Figure 5**). In *Adult2*, BMI is not correlated with age (**Figure 2**); however, titer levels and seroconversion are generally lower than in the other subpopulation (**Figure 1**). Similar to our interpretation of a saturation of an age effect, we argue that in adults older than 65 years, there is *no additional* impact of BMI on seroconversion. In comparison, the small and positive contribution of BMI to *Seroconversion* in the *Adult1* subpopulation (**Figure 5**) can be explained by the clear negative age effect (**Figure 5**) as well as a substantial correlation between age and BMI (**Figure 2**). In other words: as age, in addition to prior vaccination and baseline titer levels, accounts for much of the variation in *Seroconversion* amongst *Adult1* participants, the additional contribution of BMI is only minor.

Our results illustrate the complexity of the effects of multiple factors on the response to vaccination, their non-linear effects and interactions with each other which renders prediction of vaccine efficacy to be a continuing challenge. While our work presents a comprehensive analysis of the factors affecting seroconversion, many additional variables exist that might affect the outcome. For example, genetic makeup, pregnancy, and even the microbiome are shown to affect severity of a flu infection (Kenney *et al*, 2017; Ghedin & Schultz-Cherry, 2017; Borges *et al*, 2018), and might have to be considered in future cohort studies.

The results of our study present an opportunity to complement our ability to identify novel markers of the vaccine response: the residual of the observed and predicted *Seroconversion* discussed here can serve as a new score, i.e. a *Corrected Seroconversion* score, which simultaneously accounts for a number of known factors that are typically thought to confound the response to vaccination, such as prevaccination status, age, BMI, sex, or month of vaccination. Using the *Corrected Seroconversion* would allow identifying markers of the vaccine response *independent* of what can be explained by the participant’s specific background. This would, therefore, serve to complement the “standard” screening using the uncorrected *Seroconversion* to discover new factors impacting the response to a vaccine.

## Methods

### Data pre-processing

As part of an ongoing study by the University of Georgia, Athens (UGA), a total of 690 participants had been recruited during five seasons between 2016 and 2020 (UGA1-5). Participants received the split-inactivated influenza vaccine Fluzone by Sanofi Pasteur and provided blood samples on the day of vaccination (Day 0) (sample collected before the vaccination event) and 21/28 days post vaccination (Day 21/28). Hemagglutination inhibition assays were performed on the blood samples against each of the vaccine strains (**Figure 1**). Demographic data including age, body mass index (BMI), sex, race, comorbidities, prevaccination history, as well as month of vaccination in a flu season, vaccine dose, and baseline D0 HAI titer levels or post vaccination D21/28 titer levels were used for *Seroconversion* and *BaselineSY* predictions.

We used *Seroconversion* as log_2_-transformed ratio of HAI titer levels at day 21/28 (D21/28) and day 0 (D0) for each strain, and a composite *Seroconversion* as the sum of log_2_-transformed ratios across the strains, as proposed in a previous study (Abreu *et al*, 2020). We also log_2_-transformed age and BMI to account for their potential non-linear impact. We removed data points in which the participant had a “mixed” prevaccination status, i.e. had been vaccinated two or three years prior to the vaccination (**Supplementary Figure S1**). The remaining participants were either naive (vaccinated >3 years ago) or prevaccinated (vaccinated 1 year ago). To predict *Seroconversion*, we used the log_2_-transformed baseline D0 HAI titer levels, age, BMI, as well as the other demographic factors (variables). To predict *BaselineSY*, we used the same variables, except that the log_2_-transformed D0 HAI titer is replaced with log_2_-transformed D21/28 HAI titer (**Figure 3**). We had a total of 1,368 vaccination events across five UGA cohorts for *Seroconversion* prediction, and 672 ones across four newly assembled cohorts (repeated participants between neighboring UGA cohorts, i.e., between 1 and 2, 2 and 3, 3 and 4, and 4 and 5, respectively. They are called Baseline cohorts 1-4) for *BaselineSY* prediction.

We performed the prediction in three subpopulations separately: *Children* (<18 years), *Adult1* (18-64 years), and *Adult2* (>=65 years). We performed this split based on what is common in this field and on the fact that the high dose version of the vaccine is offered as an option to adults >=65 years only – so the effect of dose can only be modeled among *Adult2* participants.

### Predictive modeling and evaluation with an independent cohort

We first split the dataset into training and test sets. For *Seroconversion* prediction, we used UGA cohorts 1-3 and 5 as the training set and cohort 4 as the test set, as cohort 5 is a relatively biased dataset. Similarly, we used Baseline cohorts 1-2 and 4 as the training set and cohort 3 as the test set.

We then tested different machine learning algorithms in WEKA environment (Ivanciuc, 2008) (https://sourceforge.net/projects/weka/), including Random Forest and Neural Network, and selected multiple linear regression (MLR) as it had the best performance (the largest *R*^2^ for the test set). So we applied MLR to model the effect of each variable and to predict *Seroconversion* and *BaselineSY*, for individual strains as well as the composite score. To reduce redundant variables and avoid potential overfitting, we first did feature selection on the training set. We used WrapperSubsetEval attribute evaluator with the internal classifier set to be LinearRegression, and BestFirst search method, as well as a 10-fold cross-validation setting to do feature selection. Variables that were selected >=1 time out of 10 times were retained. We then developed the MLR model with the selected variables with the training set, with default settings. Finally, we evaluated the performance of the model with the test set, by both *R*^2^ and the accuracy of predicting categories (**Figure 3**).

### Estimating the relative importance of variables

We estimated the relative importance of each variable with the Leave-One-Covariate-Out (LOCO) method as described previously (Lei *et al*, 2018). Briefly, we removed one variable out at a time from the MLR model, obtained a new *R*^2^, calculated the decrease in *R*^2^ as a percentage defined as (*R*^2^_full model_ – *R*^2^_reduced model_)/*R*^2^_full model_), and used this metric as the relative importance of the variable. **Supplementary Tables S1 and S2** show the results of this evaluation.

## Data Availability

All data produced in the present study are available upon reasonable request to the authors.

https://github.com/sw5019/Fluvacc-metadata-project

## Data availability

All scripts and models were deposited on github (https://github.com/sw5019/Fluvacc-metadata-project).

## Acknowledgments

CV and TR acknowledge funding by the US National Institutes of Health (R35GM127089 and 75N93019C00052/NH/NIH HHS/United States). HC was supported in part by National Medical Research Council, Singapore (NMRC/CG/M009/2017). Cohort data was obtained from a study supported by the National Center for Advancing Translational Sciences of the National Institutes of Health under Award Number UL1TR002378. EG was supported in part by the Division of Intramural Research (DIR) of the NIAID/NIH.

The authors declare that they have no conflict of interest.

## Supplementary Notes

**Figure S1.**
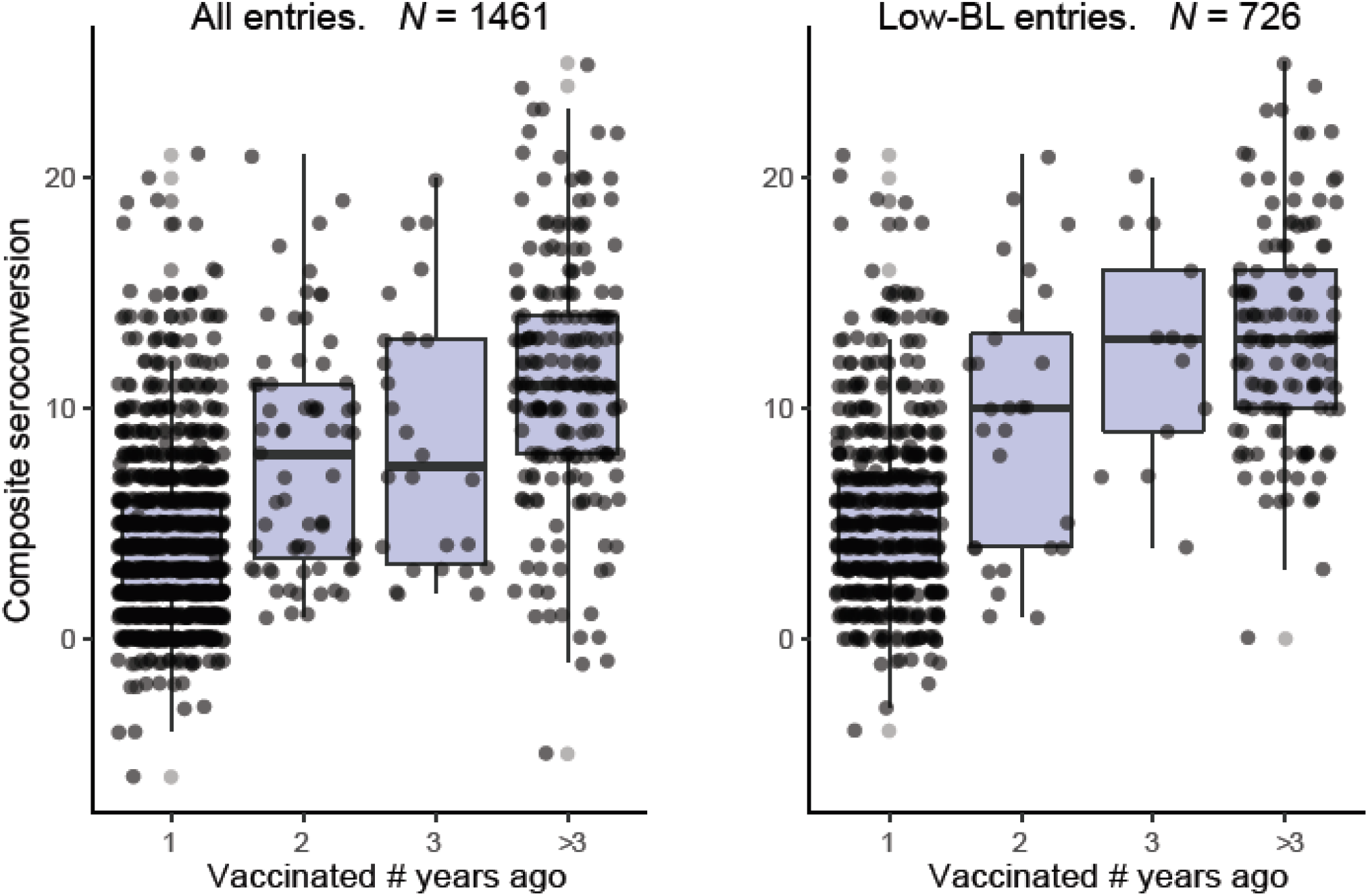
Impact of prevaccination history on seroconversion. Left panel: among all participants (*N* = 1,461); Right panel: among participants with a low baseline (seropositive strains =<2. *N* = 726).

**Figure S2.**
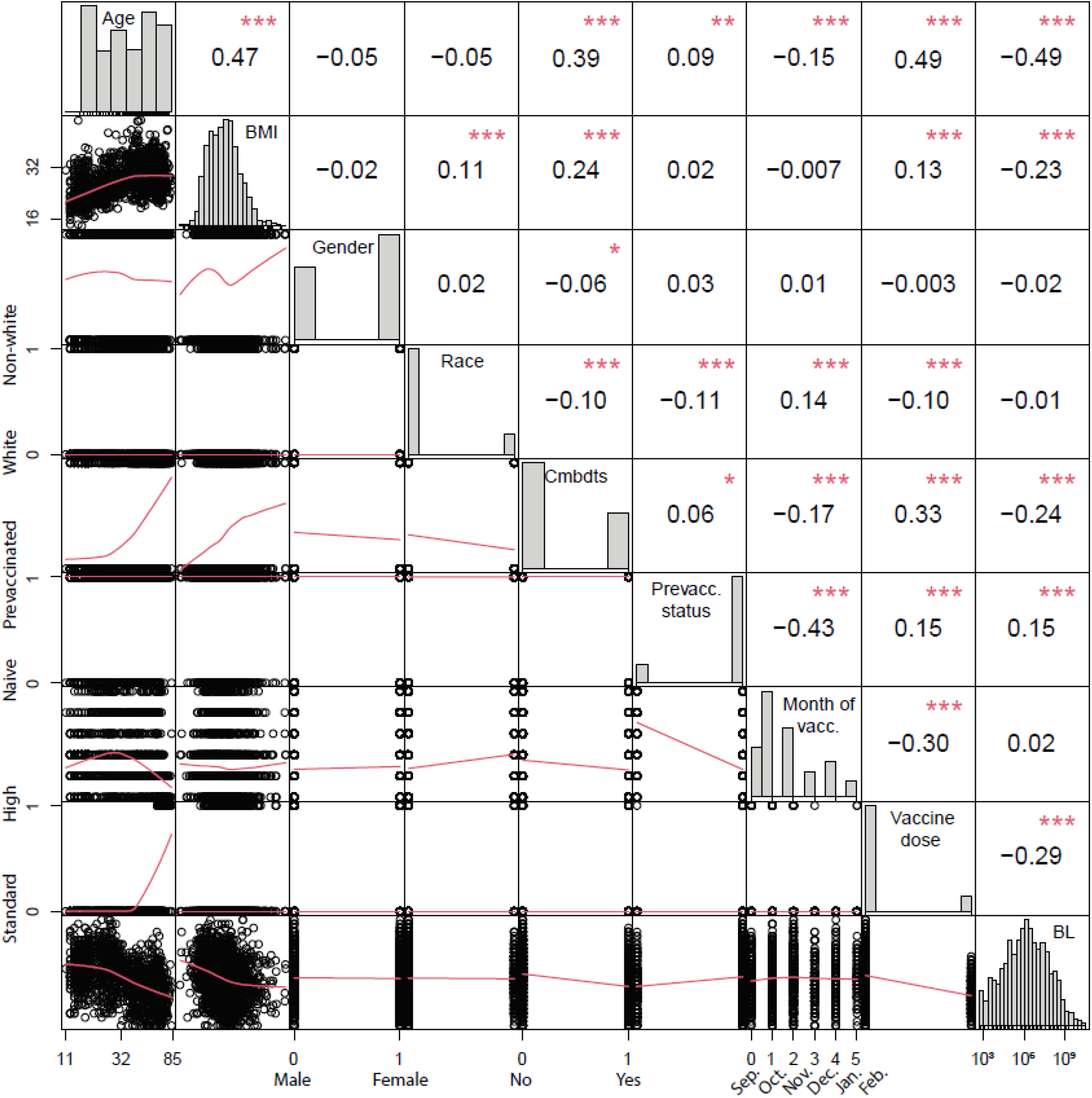
Intercorrelations between all variables. Intercorrelations between all variables that are included in the modeling. Age and BMI are log2-transformed, and the categorical variables are transformed into numeric values first, then Pearson’s correlation is calculated between the variables. *, P<0.05; **, P<0.01; ***, P<0.001. Plots along the diagonal are distributions of the variables. The red lines in the correlation plots below the diagonal are loess regression lines. Cmbdts: comorbidities; Prevacc: prevaccination; Vacc: vaccination; BL: baseline - here composite baseline.

**Figure S3.**
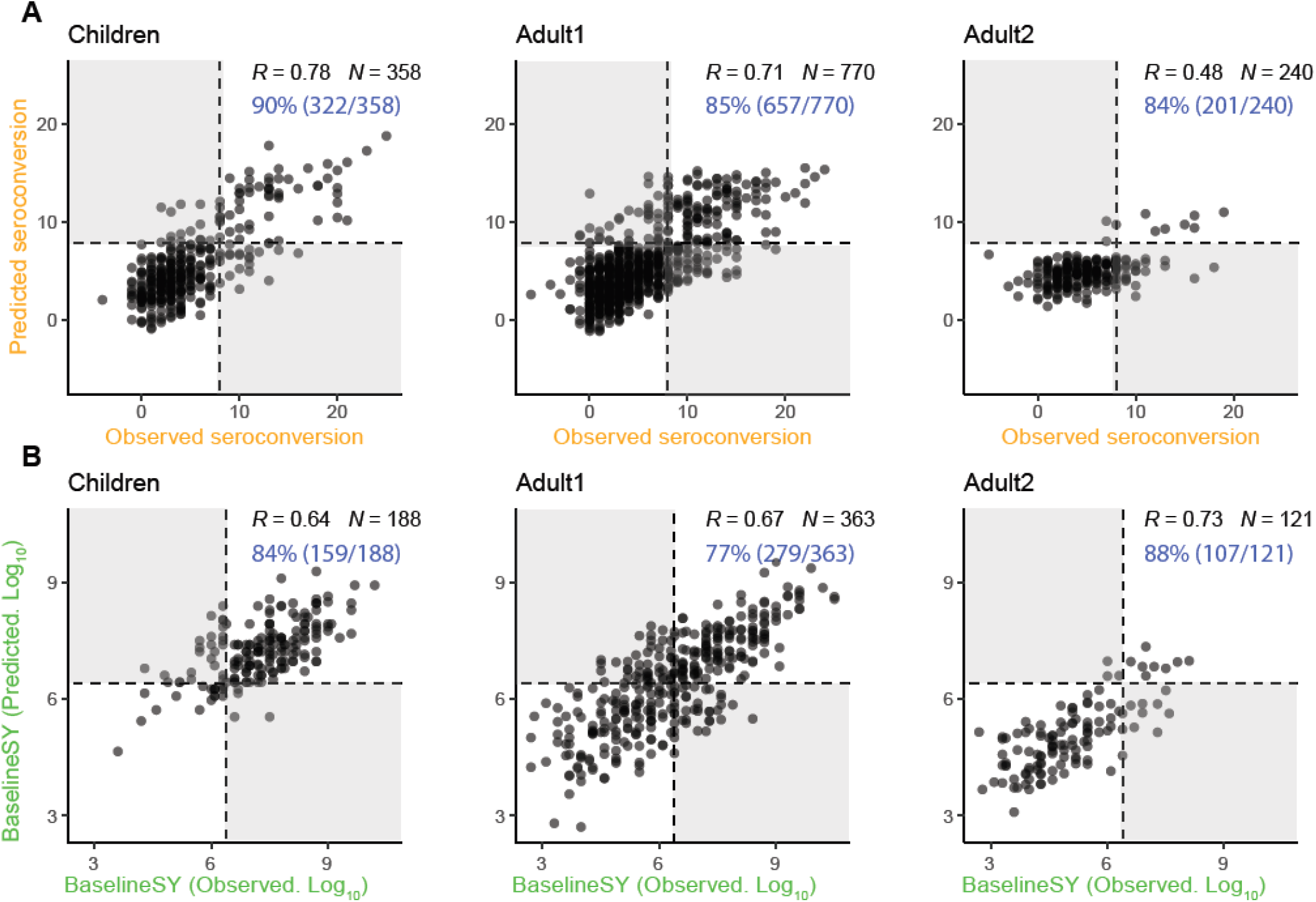
Observed vs. predicted values for *Seroconversion* and *BaselineSY*, respectively. Similar to Figure 4, only that all data points are plotted here.

**Table S1. Relative importance and coefficients of variables for *Seroconversion*** and *BaselineSY* prediction, respectively – original data.

Excel file.

**Table S2. Relative importance and coefficients of variables for *Seroconversion*** and *BaselineSY* prediction, respectively, for individual strains – original data.

Excel file.

